# Systematic prediction of mortality in trauma patients based on Arterial Blood Gas

**DOI:** 10.1101/2020.05.18.20104273

**Authors:** Zeinab Shayan, Mohammad Sabouri, Milad Shayan, Shahram Paydar

## Abstract

**Context:** – An effort to predict the final condition of patients is one of the purposes of many studies; since it enables the treatment system to provide the necessary facilities in the best possible time and prevent wasting time and energy as well as increasing patient mortality.

**Research purpose:** – This study was purposed to investigate the correlation between arterial blood gas (ABG) and patient mortality and design a system to predict the final patients’ condition.

**Method:** – In this study, a method has been proposed to identify dynamic systems to estimate the final condition of trauma patients and predict their death or survival probability during treatment or being confined in the medical center. The proposed method by using the information of patients’ arterial blood gases identifies a linear model indicating the correlation between these gases and the patients’ final condition. This method is based on system identification using ARX model simulated in MATLAB and its results are presented.

**Results:** – Data of 2802 patients (365 deaths and 2437 survivors) with an average age of 37.87 years old and GCS average of 9.27 including 470 female and 2332 male patients were studied. The designed structure was tested with 62.57% accuracy to be able to predict patient mortality. Therefore, it can be stated that the proposed method has a good accuracy in predicting the final patients’ condition based on dynamic analysis.

**Discussion and Conclusion:** – It is unavoidable mortality due to accidents and severe injuries. Also, it is important to predict the death probability based on data from the early hours of the onset of trauma in patients; since it takes time to collect the data of patient’s condition. Therefore, it is very important to find reliable methods to measure the patients’ condition and predict the mortality. The study of these methods has always been considered by physicians due to its high importance. This study has almost been able to meet physicians’ need by providing a method based on the study of dynamics and dynamic relationships discussing arterial and mortal blood gases.

## 1. Introduction

Trauma is one of the main causes of death in the world and many people including young ones are a wide range of victims in a society. Many trauma deaths occur by day 1 to 2 [1]. Researchers have always considered to be aware of trauma patients’ condition and anticipate upcoming events to be able to confront various situations ahead. Trauma surgeons are no exception. They have always been trying to find ways to determine the patients’ condition and prognosis in order to be more prepared to manage what has happened and will happen to the patient. Also, knowing the final patients’ condition before getting infected can be a key point for effective and preventive actions to change the condition. So, this has always been considered in medical researches and studied and analyzed in various aspects. However, good results have been derived from the research in this field but the research validity has always been criticized for its operationalization.

Although anticipation in primary phases is considered as a big problem [2] but different models for prediction have been studied. Many of these models used physiological and anatomical data to predict the trauma consequences including the Glasgow Coma Scale(GCS) [1], Injury Severity Score (ISS) [3-5] and its modified version [6] (New ISS) and [7] (RTS) Revised Trauma Score (RTS). Laboratory data have also been used in these models [8]. Ouellet et al. [9] studied 2269 patients and examined the value of blood gas analysis in mortality prediction in trauma patients and found that there is a significant correlation between blood gas values, especially lactate and BD levels, and mortality and length of hospital stay. In a similar study, however, Gale et al. [10] compared the value of serum lactate level and open deficiency in predicting mortality in trauma patients. They evaluated 1,829 patients and found that both lactate and BD were reported to be very high in deceased individuals, and both factors predicted premature mortality.

Aukema et al. [11] tried to validate the Thoracic Trauma Severity Score (TTSS) by referring to the 712 data banks of patients to investigate the side effects of this trauma and levels of patient injury. They stated that patients who died from chest injuries and their side effects, including ARDS, clearly had a higher TTSS than patients who died of other injuries. They stated that the score could predict mortality as well as ARDS.

In a study, Ahun et al. [12] studied GCS, age, and arterial blood pressure as a physiological score for predicting mortality in emergency trauma patients. They stated that this scale, along with other scales such as TRISS and ISS, can predict mortality in high power and they hoped that this could help manage trauma patients.

In this study, it is tried to provide an approach based on the dynamic study of a structure to find the correlation between arterial blood gases and trauma patients’ mortality and a method to predict the patients’ mortality over these gases. This research is organized as follows. In section 2, it is presented a proposed method and mathematics involving this method and the process. The results of the performed simulations and review are presented in section 3. Finally, section 4 concludes the conducted research.

## 2. Method

By collecting data on the arterial blood gases of PH, *PCO*_2_*, PO*_2_, BE^1^ and *HCO*_3_ related to 2802 patients, it was tried to estimate their mortality by system identification. Based on the data of these 5 inputs, the survived and died patients were labeled 1 and 0 respectively and 365 deaths and 2,437 survivors were stored as output. To examine this data, two datasets were developed as a training and test dataset of 2142 and 660 patients respectively. Totally, this study is involved 470 female and 2332 male patients in an average age of 37.87 years old and an average GCS of 9.27. Details of the number of male and female alive and dead in training and test data set are presented in Table 1.

**Table 1.** Collected data

|  | Total Data | Total Number of Deaths (0) | Total Number of Survivors (1) |
| --- | --- | --- | --- |
| **Training Data** | 2142 | 294<br>(49 females, 245 males) | 1848<br>(295 females, 1553 males) |
| **Test Data** | 660 | 71<br>(14 females, 57 males) | 589<br>(112 females, 477 males) |
| **Total Data** | 2802 | 365 | 2437 |

### 2.1. System identification

Unlike methods such as neural networks, system identification is a model-based approach using mathematical input-output data to provide a mathematical model for the system [13-14] indicating system dynamics. Linear and nonlinear models can be used to obtain model parameters using data.

The proposed model used in this paper is ARX model (linear model) to predict mortality that the system is identified by determining input u(t) and output y(t) and considering the noise e(t). Figure 1 and Equations (1) to (7) shows a block diagram and equations of this model respectively.

**Figure 1.**
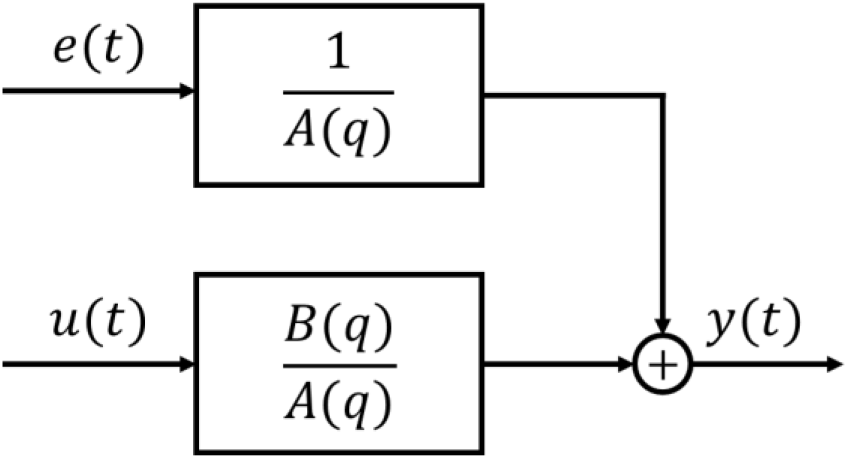
Block diagram of ARX model

For a five-input single-output system, output y(t) is a one-dimensional vector, input u(t) is a five-dimensional vector, e(t) is a noise vector, A(q) is a 1×1 matrix, and B (q) is a 1×5 matrix.

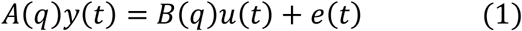

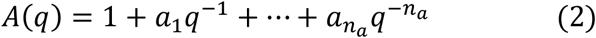

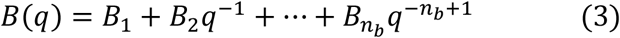

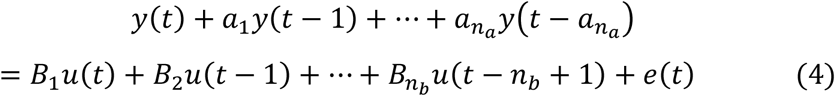

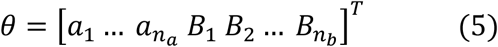

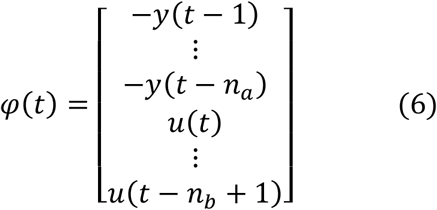

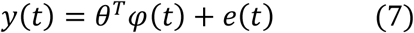

After system identification, that is, determining the parameters of the system (θ) and having an input, we can estimate the output using Equation 7.

To use this model, 2142 data were first selected, including output vector and input five, for training and system identification. Using system identification toolbox in MATLAB, an ARX model with *n_a_ =* 5 and *n_b_ =* 1 (for each input) was obtained for the data to determine the parameters’ vector, θ (Figure 2 (a)). After obtaining the parameters, input and white noise with a standard deviation of 0.01 were applied to the system and the output of ARX model was obtained based on Equation (7). This data is entered into a comparator to convert *y_ARX_* to 0 and 1; If *y_ARX_* is more and less than *y_ARX_* average, output is 1 and 0, respectively (for *y_ARX_* output, the average was obtained 0.86). In this case, the estimated output vector of whole system (*ŷ*(*t*)) is obtained (Figure 2 (b)).

**Figure 2.**
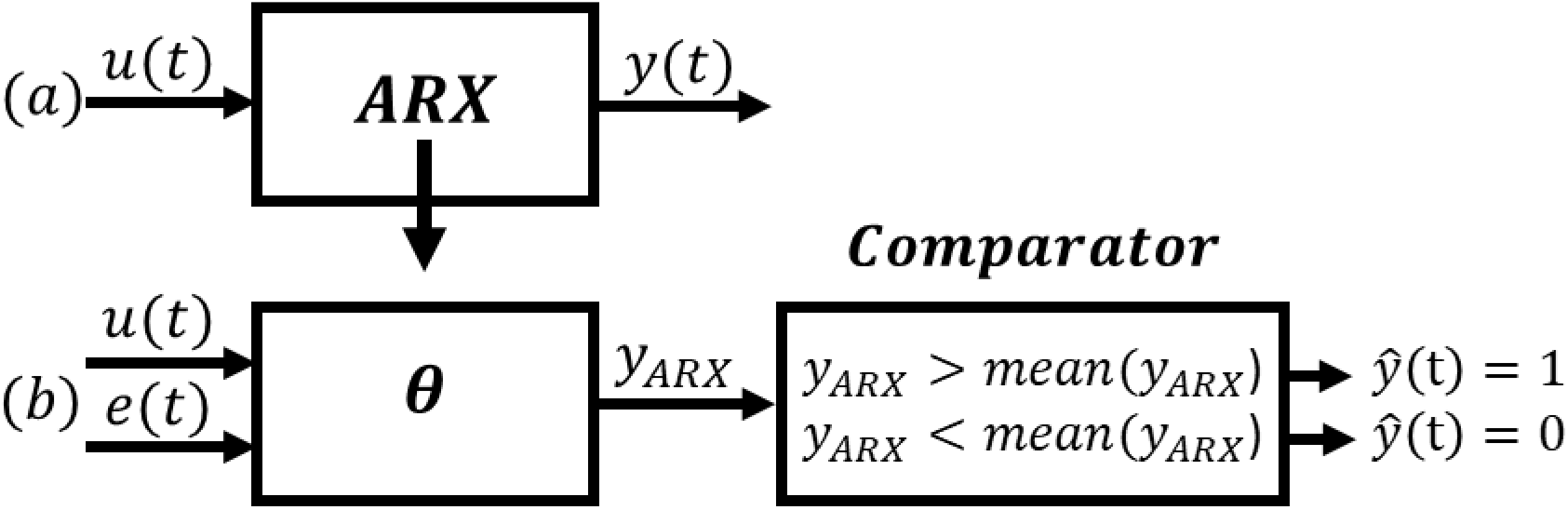
The process of obtaining final output of system: (a) ARX model parameter by training data (b) Output estimation by obtained parameters

The MSE^2^ was used to verify the data model accuracy which compares the estimated output *ŷ*(*t*) and actual output *y(t)* and includes:

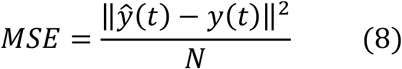

where N is number of samples and ‖ . ‖ is 2 norm vector. Therefore, percentage of the obtained model accuracy will be equal to:

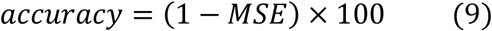

After obtaining the estimated training data and checking its accuracy, new data are applied to test the system (Figure 2 (b)).

## 3. Result and Discussion

When 2142 training data were applied to the system, the parameters of ARX model were obtained and the model was identified as (10):

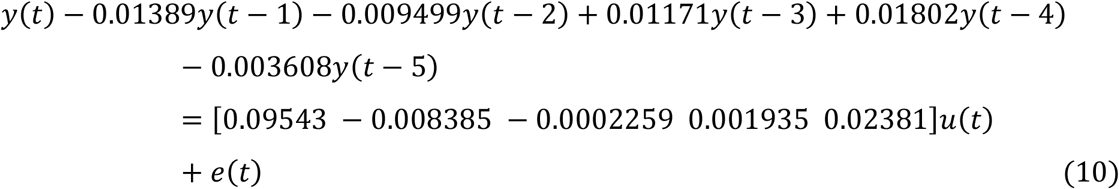

In Figure 3, the output of ARX model after applying training input, the estimated output and the actual output is shown for 50 data.

**Figure 3.**
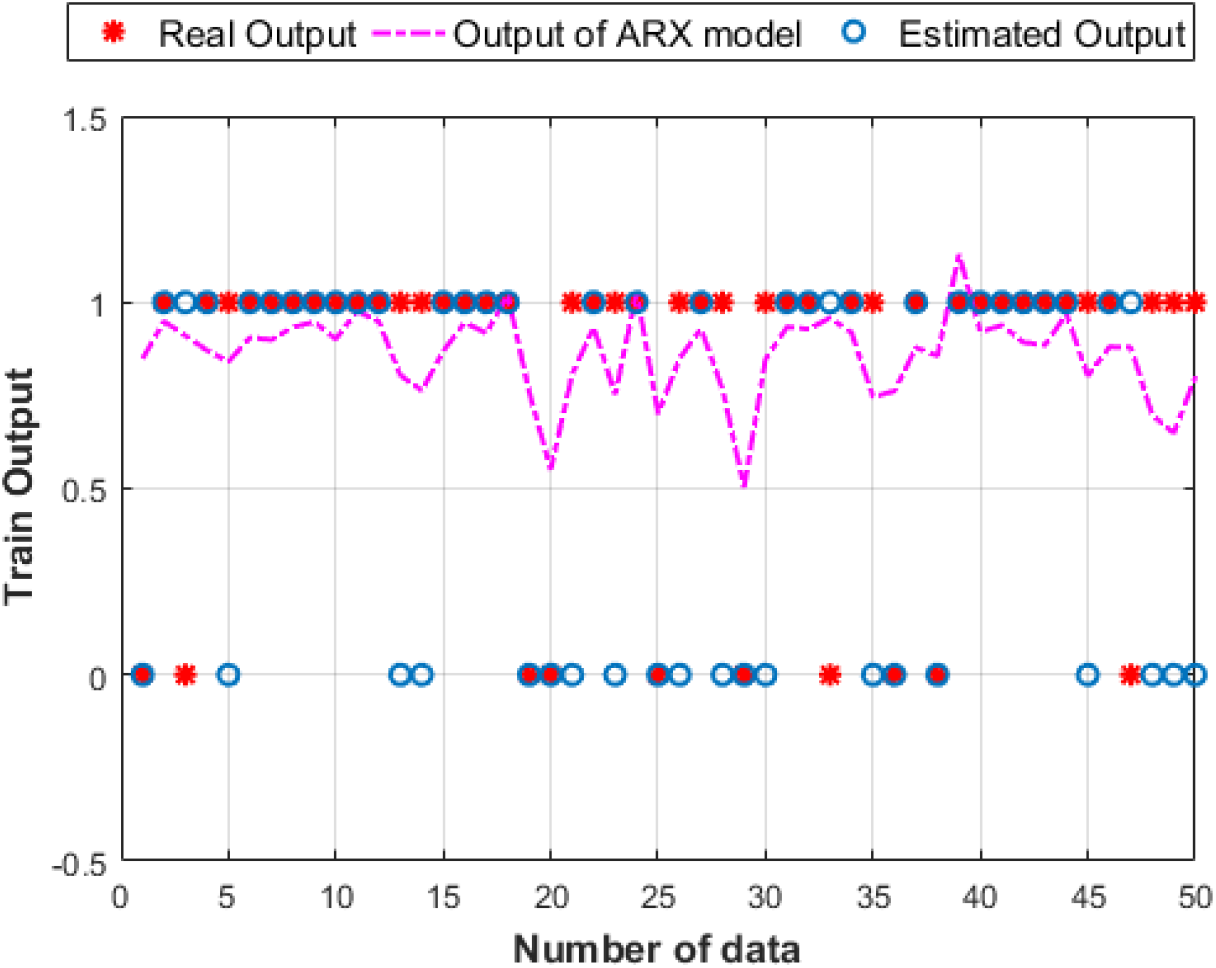
ARX model’s output, estimated and actual output of 50 training data

As shown in Figure 3, due to continuous data, ARX model’s output entered to comparator, so it is converted to discrete data 0 and 1 to estimate output. “*” is actual output and “o” is estimated output. As it is clearly observed, some of estimated outputs did not follow the actual output indicating an error in designed system and depends on the accuracy of system.

According to estimated output, the accuracy of 2142 relational training data from (9) was obtained 62.55%. Out of this, 1848 and 294 data are related to survivors and deaths respectively. Designed system identified 1,135 alive and 204 dead people, with an accuracy of 61.41% and 69.38%, respectively.

To ensure the proper functioning of designed system in estimating the mortality rate, 660 new data were applied to system as test data and the accuracy of test data was obtained 62.57%. Out of 660 test data, there are 589 survived and 71 dead individuals; however, system estimated 372 survivors and 43 deaths. Therefore, the estimated accuracy of survivors and deaths for test data is 63.15% and 60%, respectively. Table 2 presents the accuracy of training and test data.

**Table 2.** Train and Test Accuracy

|  | **Total**<br>**Number**<br>**of**<br>**Survivors**<br>**(1)** | **Number**<br>**of True**<br>**Estimated**<br>**Survivors** | **Survivors'**<br>**Accuracy** | **Total**<br>**Number**<br>**of**<br>**Deaths**<br>**(0)** | **Number**<br>**of True**<br>**Estimated**<br>**Deaths** | **Deaths'**<br>**Accuracy** |
| --- | --- | --- | --- | --- | --- | --- |
| **Train**<br>**(62.55%)** | 1848 | 1135 | 61.41% | 294 | 204 | 69.38% |
| **Test**<br>**(62.57%)** | 589 | 372 | 63.15% | 71 | 43 | 60% |

In designed system, threshold value considered in comparator is a parameter that is able to change and alter the accuracy of system. Therefore, training and test data were applied to the system again, but the data were compared with *y_ARX_ =* 0.8 rather than *y_ARX_ =* 0.86. In this case, the accuracy of test data was obtained 80.6% indicating that the accuracy has increased by decreasing threshold value. However, the estimated accuracy of the number of survivors and deaths in this case was 85.5% and 40% respectively. It shows that despite the increased accuracy of system and survivors, the estimating accuracy of deaths is decreased. System estimated the deaths in less accuracy because there are heterogeneous data and the number of survivors are more than deaths causing to make a more successful system in training survivors. Therefore, the system must have high overall accuracy and the estimated accuracy of survivors and deaths is high, as well. Thus, it is purposed to design a model using the method of system identification in future which can estimate the status of survivors and deaths in high accuracy despite data heterogeneity.

## 4. Conclusion

According to the results and accuracy obtained from the designed structure, it seems that this structure based on the dynamics involving in data and the systematic review of data has been able to find an effective correlation between arterial blood gases and mortality in trauma patients. Based on the accuracy obtained (62.57%) on the test data firstly given to the structure, it can be stated that the prediction by this accuracy is highly significant. This can be used as a new method rather than traditional one in predicting mortality and measuring the final condition of patients.

It is notable that this research is being completed and upgraded; also, the results presented in this study are preliminary.

## Data Availability

The data that support the findings of the present study are available from the Rajaee (Emtiaz) Hospital and are not publicly available. The anonymized dataset used for the present research is however available from the corresponding author on reasonable request and with permission of both Trauma Research Center and Rajaee (Emtiaz) Hospital of Shiraz.

## Acknowledgments

The authors would like to thank the members of Shiraz Trauma Research Center of Shahid Rajaee (Emtiaz) Hospital members, especially Dr. Shahram Bolandparvaz, head of Trauma Research Center, for providing this research opportunity, and also Dr. HamidReza Abbasi, deputy of Trauma Research Center for his cooperation and supporting in this research.

## Authors’ contributions

ZSH and MS and MSH contributed to the study conception, implementation of the study, design, interpretation and critical was involved in drafting of the manuscript. SHP contributed to the conception and design of data and revising the manuscript. All authors have read and approved the manuscript, and ensure that this is the case.

## Ethics approval and consent to participate

This study was conducted according to the principles expressed in the Trauma Research Center of Shahid Rajaee (Emtiaz) Hospital of Shiraz and approved by the local ethics committee of Shiraz University of Medical Sciences by the code IR.SUMS.REC.1397.719. Based on the approval of Ethics Committee, all informations were collected only by patient’s code and their identity was not disclosed. Patient’s information was in private.

## Footnotes

1 Base Excess

2 Mean square error

